# Noise-Assisted MEMD-Based Phase-Connectivity Analysis to Personalize Closed-Loop DBS Therapy in Epilepsy Patients

**DOI:** 10.1101/19006908

**Authors:** Sina Farahmand, Tiwalade Sobayo, David J. Mogul

## Abstract

Deep brain stimulation (DBS) is a treatment that has been explored for controlling seizures in patients with intractable epilepsy. Many clinical and pre-clinical studies using DBS therapy determine stimulation parameters through trial and error. The same stimulation parameters are often applied to the whole cohort, which consequently produces mixed results of responders and non-responders. In this paper, an adaptive non-linear analytical methodology is proposed to extract stimulation frequency and location(s) from endogenous brain dynamics of epilepsy patients, using phase-synchrony and phase-connectivity analysis, as seizures evolve. The proposed analytical method was applied to seizures recorded using depth electrodes implanted in hippocampus and amygdala in three patients. A reduction in phase-synchrony was observed in all patients around seizure onset. However, phase-synchrony started to gradually increase from mid-ictal and achieved its maximum level at seizure termination. This result suggests that hyper-synchronization of the epileptic network may be a crucial mechanism by which the brain naturally terminates seizure. Stimulation frequency and locations that matched the network phase-synchrony at seizure termination were extracted using phase-connectivity analysis. One patient with temporal lobe epilepsy (TLE) had a stimulation frequency ∼15 Hz with the stimulation locations confined to the hippocampus. The other two patients with extra-temporal lobe epilepsy (ETE) had stimulation frequency ∼90 Hz with at least one stimulation location outside of hippocampus. These results suggest that DBS parameters should vary based on the patient’s underlying pathology. The proposed methodology provides an algorithm for tuning DBS parameters for individual patients in an effort to increase the clinical efficacy of the therapy.

## I. Introduction

Epilepsy is a chronic disorder of the brain characterized by recurrent, spontaneous, and abnormal brain activity called epileptic seizures [1]. Epilepsy patients may undergo unusual behavior, sensations, or loss of consciousness during seizures. Approximately, 70 million people worldwide are living with epilepsy, in which about one-third of them do not respond to antiepileptic medications or become drug-resistant [2], [3].

Epilepsy surgery is an alternative treatment for these drug-refractory patients, in which regions of the brain responsible for generating seizures are resected [4], [5]. However, only patients with well-localized epileptogenic zones in an area out of the eloquent cortex are surgical candidates. For drug-refractory epilepsy patients who do not benefit from resecting surgery, an alternative therapeutic approach is needed.

Deep brain stimulation (DBS) has been widely evaluated for controlling intractable seizures [6]-[8]. DBS involves delivering electrical impulses to electrodes implanted within specific areas of the brain. DBS has the capacity to modulate brain activity in a controlled and reversible manner [9]-[11]. Although the therapeutic mechanisms underlying DBS are not well understood, it is the only effective therapy for many people suffering from refractory neurological disorders.

Open-loop DBS and closed-loop DBS are the two, primary approaches that have been utilized in order to control seizures over the last decade [12], [13]. In open-loop DBS, electrical stimulation is delivered to the target location(s) in a pre-programmed manner and independent of a patient’s brain dynamics and clinical symptoms [14], [15]. That is, there is no embedded unit in open-loop stimulation devices to record and analyze endogenous brain activities of patients and adjust the stimulation parameters accordingly in order to improve seizure control. In open-loop DBS systems, electrical stimulation is applied either continuously or intermittently based on a clock-determined cycle [16]. SANTE (Stimulation of the Anterior Nuclei of Thalamus for Epilepsy) study using Medtronic DBS device, Activa PC, (Medtronic, Minneapolis, MN, USA) was performed in an open-loop manner to reduce the frequency of seizures in epilepsy patients [17]. This clinical study reported that those patients who responded to the therapeutic thalamic stimulation experienced approximately 41% reduction in their seizure frequency, over 4 months of DBS, relative to baseline seizure frequency prior to the start of stimulation [17].

Alternatively, in an ideal closed-loop DBS, stimulation parameters are tuned based on a patient’s brain activity and clinical symptoms. Electrical stimulation is triggered in response to seizure detection [18]. NeuroPace Responsive Neurostimulator System (RNS), (NeuroPace, Mountain View, CA, USA), has been used in a closed-loop manner to reduce the frequency of seizures in epilepsy patients [19]. The RNS delivers DBS upon seizure detection or imminent onset; however, stimulation parameters are pre-determined and independent of a patient’s endogenous brain dynamics. The RNS clinical trials reported an approximately 50% reduction in seizure frequency in about 54% of patients [19].

In most of the clinical and pre-clinical studies using DBS therapy, electrical stimulation parameters such as frequency and amplitude are derived based on trial and error [20], [21]. Alternatively, a therapeutic DBS protocol based on real-time seizure dynamics measured and evaluated prior to the next seizure could potentially improve clinical efficacy and may potentially result in personalizing and optimizing DBS therapy for individual epilepsy patients. In our previous study, we validated the effectiveness of employing this approach in terminating seizures in a freely moving chronic rat model of limbic epilepsy [22].

It has been widely shown that synchrony among neuronal oscillators could be altered in epilepsy, which itself may underlie seizure onset [23]-[25]. Therefore, evaluating the synchrony dynamics within an epileptic network may provide essential insight into seizure dynamics as they evolve. However, identifying the synchrony level between non-linear, non-stationary neuronal oscillators extracted from intracranial electroencephalographic (iEEG) data represents a challenge. Furthermore, it has been shown that phase synchronization can be detected among non-linear, coupled oscillators even if their amplitudes are not synchronized [26]. Therefore, it is critical to utilize non-linear methods to measure instantaneous phase-synchrony. In our previous study, an analytical methodology based on noise-assisted multivariate empirical mode decomposition (NA-MEMD) was utilized to quantitatively evaluate the phase-synchrony dynamics in patients with temporal and frontal lobe epilepsy [27]. In this study, a reduction in the phase-synchrony was observed around seizure onset. However, the network phase-synchrony started to gradually increase towards seizure termination and achieved its maximum at seizure offset. The synchrony dynamics observed in this study were consistent with several previous studies in the literature [28]-[30]. The phase-synchronization observed as seizures naturally end may be an endogenous mechanism for seizure termination.

This study reports on developing an adaptive, non-linear analytical methodology that merges the phase-synchrony and phase-connectivity analyses in order to extract frequency and locations of stimulation that match the synchronized network-dynamics at seizure termination.

In this study, a DBS protocol using the extracted frequency and locations matching the hyper-synchronization dynamics observed in epilepsy patients as seizures naturally terminate is proposed that could be used to potentially revert the network desynchronization observed around seizure onset in order to immediately terminate pathological electrical activity.

The rest of the paper is organized as follows. In section II, specifications of the adopted iEEG dataset, along with the framework of the proposed phase-connectivity analysis are described. Next, experimental results of extracting stimulation parameters in all patients using phase-connectivity analysis are presented in section III. Finally, section IV discusses the results and provides conclusions to this paper.

## II. Dataset and Methods

The conceptual framework of the proposed instantaneous phase-synchrony analysis is shown in Fig. 1(a). Multi-channel iEEG data, recorded from different brain regions of epilepsy patients using implanted depth electrodes, were used as input to the analytical signal processing procedure. A set of finite neuronal oscillators, IMFs, were extracted using NA-MEMD from raw iEEG data. Next, Hilbert transform was performed on the decomposed neuronal oscillators in order to measure their instantaneous phases and frequencies. A combination of mean-phase coherence analysis and eigenvalue decomposition technique was then employed to evaluate the phase-synchrony dynamics among neuronal oscillators as seizures evolved.

**Fig. 1.**
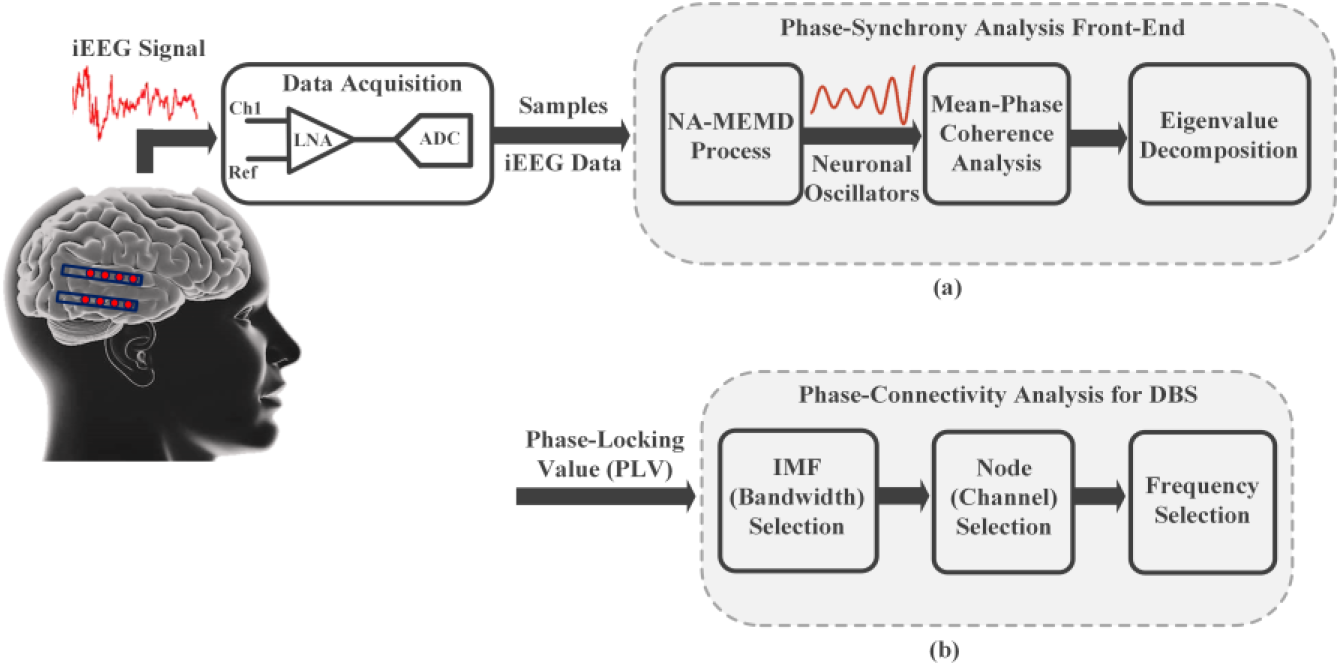
Conceptual front-end of the proposed (a) phase-synchrony analysis, and (b) phase connectivity analysis.

Fig. 1(b) illustrates the framework of the proposed phase-connectivity analysis among neuronal oscillators to extract the frequency and locations of stimulation. This process involved selecting IMF (bandwidth of interest), nodes (locations of stimulation), and finally frequency of stimulation. It should be noted that the aforementioned parameters were extracted to match the maximum level of phase-synchronization observed at seizure termination.

### A. Patients and iEEG Dataset

In this study, multi-channel iEEG data recorded from depth electrodes implanted in three de-identified epilepsy patients were retrieved from the online International Epilepsy Electrophysiology (IEEG) portal [31]. Two patients were diagnosed with extratemporal lobe epilepsy (ETE) with left temporal onset, while the third patient had temporal lobe epilepsy (TLE) with a right temporal seizure focus. All patients were adults undergoing long-term iEEG monitoring in preparation for the pre-surgical evaluation including seizure onset zone (SOZ) localization of their medically refractory epilepsy. Epileptic SOZs and timing of seizures, seizure onset and termination, were clinically determined and annotated by the treating physicians at Mayo Clinic via visual inspection of clear ictal discharges. Further clinical information regarding the analyzed patients as well as their access IDs in the IEEG portal are provided in Table I. The iEEG data were recorded from the patients using depth electrodes with 4 or 8 recording sites, a 500 Hz sampling rate, and a 1-150 Hz bandpass filtering. All epilepsy patients were implanted with two depth electrodes in both amygdala and hippocampal regions of the brain. A digital 60 Hz notch filter was used to eliminate line noise.

**TABLE I.**
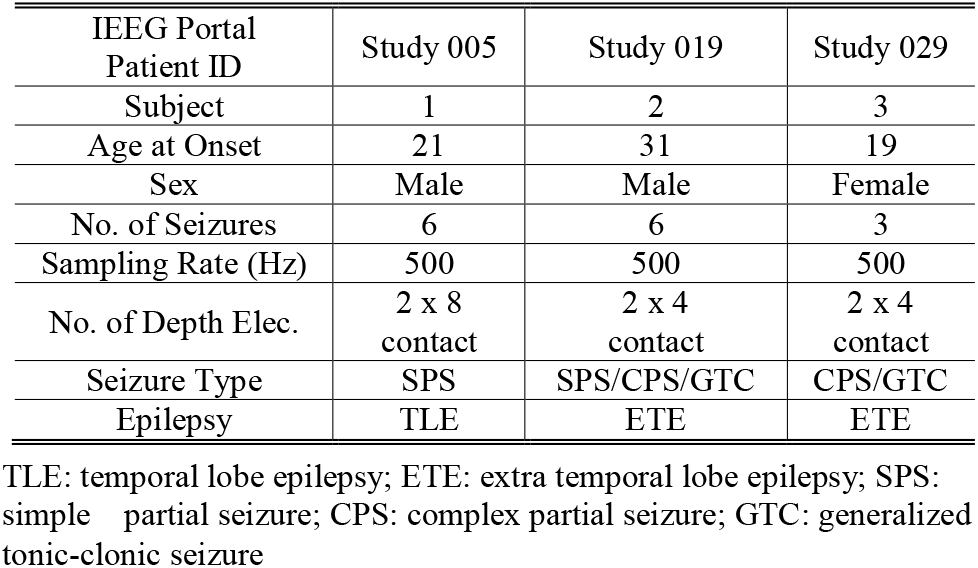
PATIENT CLINICAL DATA AND SIGNAL ACQUISITION INFORMATION

### B. Noise-Assisted Multivariate EMD (NA-MEMD)

EMD is an adaptive, data-driven method of decomposing signals into a set of finite and nearly orthogonal oscillators, called intrinsic mode functions (IMFs) [32]. The decomposed IMFs together form the underlying oscillations within a time series. That is, IMFs are truly determined from the dynamics of original signals considering the nature of their underlying components. Therefore, EMD is an appropriate method for the time-frequency analysis of non-linear, non-stationary electrophysiological signals [33]. However, EMD of multi-channel data can result in mode-aliasing and mode-misalignment [34]. An advanced version of the EMD method called NA-MEMD was introduced to overcome the aforementioned problems which is described in detail in [35]. The original signal is described by the following equation at the end of the decomposition.

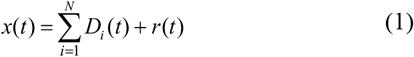

where *N* is the number of the extracted IMFs and *r(t)* is the residue. The residue corresponds to a signal whose projections do not contain sufficient extrema to comprise a multi-variate envelope. The lower-index IMFs correspond to fast oscillation modes of the original signal while the extracted higher-index IMFs denote the slower oscillation modes.

In this study, NA-MEMD was performed on iEEG data segments to obtain its corresponding IMFs. The instantaneous phases of IMFs were measured using the Hilbert transform. The NA-MEMD on a 1-s ictal data-segment along with the time-frequency representation of its extracted IMFs are shown in Fig. 2. For simplicity, only the top six extracted IMFs are illustrated in Fig. 2. The in-depth rationale for employing NA-MEMD prior to the determination of the instantaneous phase information are discussed in the next section.

**Fig. 2.**
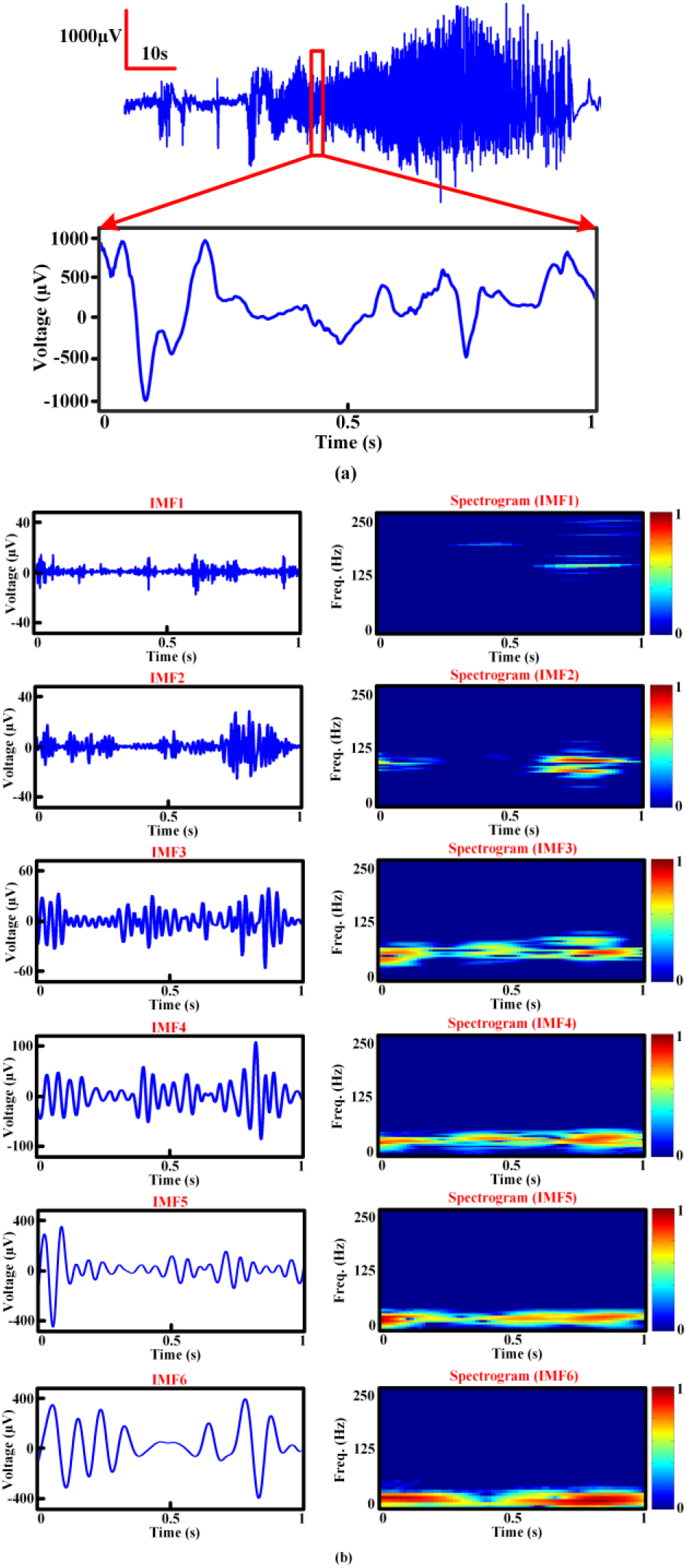
NA-MEMD process on an iEEG recording. (a) 1.5-min iEEG signal along with a magnified view of its 1-s ictal period utilized for the NA-MEMD process. (b) The top six IMFs decomposed from the 1-s, ictal period along with their time-frequency representations.

### C. Hilbert Analytic Signal Method

Hilbert analytic signal method has been widely utilized to extract instantaneous phase and frequency information from narrowband signals [36]. Details regarding using the Hilbert transform to extract instantaneous phase points and frequency information from a time series are described in [36].

In order to extract unambiguous phase information, the phase space trace of an analytic signal should possess a single center of rotation. Multi-component or wideband signals yield trajectories in the complex plane with multiple centers of rotation. Therefore, it is essential to individually extract the underlying components of a time series, contributing to the multiple centers of rotation, in order to reach an unambiguous phase reading. NA-MEMD method, described in the previous section, provides a proper instantaneous phase determination by separating a wideband signal into its underlying oscillatory components or IMFs. Each one of these IMFs results in a complex plane with a single center of rotation. In this study, NA-MEMD of iEEG signals were performed for each 1-s segment of data prior to the use of the Hilbert transform to measure the instantaneous phase points.

### D. Mean-Phase Coherence Analysis

In order to determine the strength of phase relationship among the extracted IMFs, the instantaneous phase points of each IMF was represented as a row vector and stacked on top of each other to form an *N* ***×*** *M* matrix, in which *N* is the total number of IMFs decomposed from all channels within a 1-s data segment and *M* denotes the total length of the time series. Next, the bivariate mean-phase coherence matrix, *R*_*N×N*_, was calculated using the following equation [37].

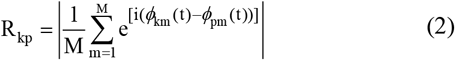

in which *i* is equal to √-1, *M* is the total number of samples within the data segment, and *Ø(t)* is the instantaneous phase of the analytic signal pair indicated by the subscripts *k* and *p*. Subscripts *k* and *p* iterate from 1 to *N* and all the values in the *R*_*N×N*_ are between zero and one.

### E. Eigenvalue Decomposition and Synchrony Analysis

Eigenvalue decomposition of the square, bivariate mean-phase coherence matrix was carried out in order to achieve a multi-variate measure for capturing phase-synchrony among all the extracted neuronal oscillators. All the eigenvalues were sorted in ascending order to construct an eigenvalue spectrum. Each eigenvalue indicates how strongly oscillators are phase-correlated in the direction of its associated eigenvector.

The eigenvalue decomposition was performed by solving *R*_*N×N*_*v*_*i*_ *= λ*_*i*_*v*_*i*_, where *λ*_*i*_ and *v*_*i*_ are the obtained eigenvalues and their corresponding eigenvectors, respectively. It is important to note that all the *N* obtained eigenvalues were real with their sum equal to the total number of the IMFs, as the *R*_*N×N*_ matrix was square symmetric. Therefore, any decrease in one of the eigenvalues must be compensated by an increase in the other eigenvalues in order to keep the sum constant. This is the main idea underlying capturing the phase-synchrony between the extracted oscillators. Due to the ascending order between the eigenvalues, a small change in the value of a few higher-index or larger eigenvalues causes a relatively large dynamic change in the value of the majority of lower-index eigenvalues in order to compensate it. Hence, the temporal changes of phase-synchrony are captured with higher sensitivity by focusing on the high percentage of lower-index eigenvalues. The average value of the first 60% lower-index eigenvalues, *meanλ*_*1:60%*_, was computed for each 1-s segment of data to quantify the temporal evolution of phase-synchrony levels.

Computational modeling was described in our previous study to quantify and compare *meanλ*_*1:60%*_ value among three epileptic networks with different phase-synchrony level [27]. Simulation results demonstrated that as the phase-synchrony level increases between neuronal oscillators from its minimum to its maximum, the *meanλ*_*1:60%*_ value decreases accordingly. Furthermore, the normalized *meanλ*_*1:60%*_ values were measured to quantify phase-synchrony levels as seizures evolve in order to magnify the small relative changes of the *meanλ*_*1:60%*_ over time. It should be noted that phase-synchrony was observed to increase from mid-ictal towards seizure end and achieved its maximum level at seizure termination in all of the analyzed epilepsy patients [27].

### F. IMF (Bandwidth) Selection for DBS

A 10-s window leading up to the seizure termination was selected for phase-connectivity analysis in order to provide adequate timing for evaluating changes in phase relationships. The main purpose of the IMF selection process was to identify which of the extracted IMFs mostly contributed to the hyper-synchronization observed at seizure termination.

Fig. 3 illustrates how phase-connectivity of the extracted IMFs were evaluated. This example displays the comparison between two different IMFs in order to select the one that has higher phase-synchronization within the termination window. The same index IMFs from five iEEG channels were grouped to model the phase-connectivity analysis. Fig. 3(a) illustrates the fourth neuronal oscillators (IMF4) decomposed from five different iEEG channels within a 10-s window leading up to seizure termination. The bivariate phase-locking value, *PLV*, was then calculated in each 1-s segment for every unique pairs of channels. The *PLV* value was measured according to the following equation [38].

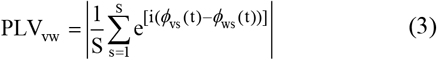

in which *i* is equal to √-1, *S* is the total number of samples within the data segment, and *ϕ(t)* is the instantaneous phase of the IMF extracted from channels indicated by subscripts *v* and *w. PLV* values can be between zero and one. Circles in Fig. 3(a) represents iEEG channels and *PLV* values indicate the strength of phase-connection between a pair of channels. For simplicity, only the *PLV* values of the connections forming the outer edge of the diagram are displayed for the first and last 1-s segments. The average *PLV* value among all connections, *M-PLV*, was then calculated for each 1-s segment. Finally, the average *M-PLV* values were measured for the entire 10-s termination window, which is defined by *TM-PLV* value. Fig. 3(b) shows the aforementioned steps on the fifth IMFs (IMF5) extracted from the same five iEEG channels within the 10-s termination window.

**Fig. 3.**
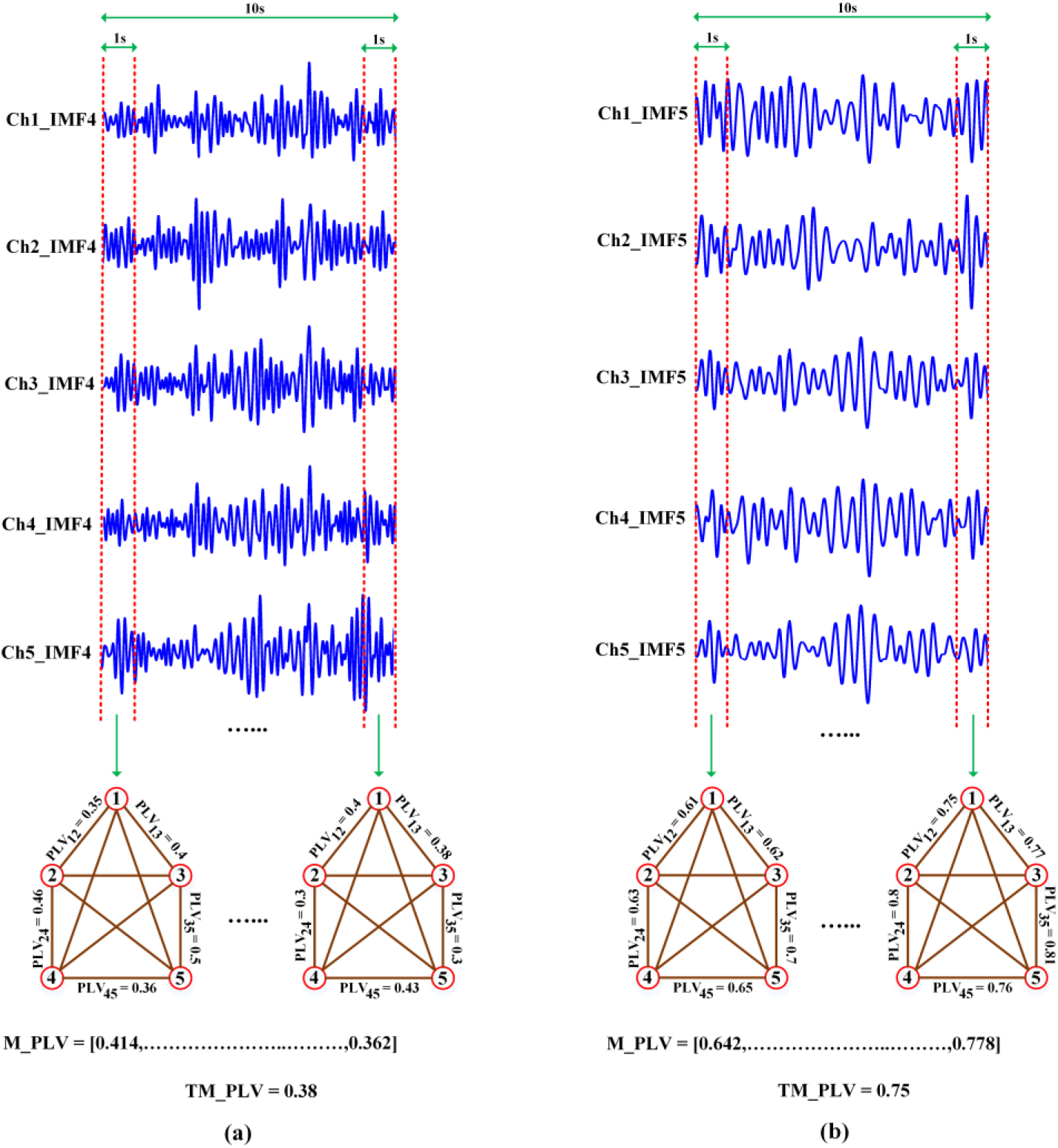
Selection of IMF (bandwidth of interest) for DBS. (a) A set of the fourth neuronal oscillators (IMF4) decomposed from five different iEEG channels within a 10-s window leading up to seizure termination. Bivariate *PLV* values were measured for every unique pairs of channels in each 1-s data segment. The average *PLV* value across the whole network, *M-PLV*, was calculated for every 1-s and its temporal average across the 10-s window, *TM-PLV*, was utilized as a metric to assess the network-level of phase-connectivity within each IMF. (b) A set of the fifth oscillators (IMF5) extracted from the same five channels within the 10-s termination window. The IMF index with the largest *TM-PLV* value was selected as the one with the highest contribution to the hyper-synchronization observed at seizure termination. For simplicity, only the PLV values of the connections forming the outer edge of the diagram are displayed for the first and last 1-s segments.

As it is shown in Fig. 3, neuronal oscillators within IMF5 resulted in a higher *TM-PLV* value relative to the ones in IMF4. Therefore, it contributes more into increasing the phase-synchrony level at termination window. The IMF index with the largest *TM-PLV* value was selected as the one with the highest contribution to the hyper-synchronization observed at seizure termination. It should be noted that the frequency bandwidth of the selected IMF was considered as the tuning range for the frequency of stimulation.

### G. Node (Channel) Selection for DBS

Once the IMF of interest is identified, the next step is to determine which iEEG channels would be the best candidates for applying DBS. First, the *PLV* values were calculated in each 1-s segment for every unique pairs of channels, within the selected IMF, for the entire 10-s termination-window. Next, the temporal average of *PLV* values was measured over the 10-s termination-window for each unique pair. This step results in a *L* ***×*** *1* array which *L* is defined as follows:

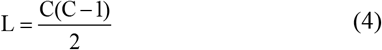

in which *C* is the total number of nodes or iEEG channels. Finally, nodes that mostly contributed to the hyper-synchrony observed at seizure termination-window, within the selected IMF, were determined by the following steps:

1. *TM-PLV* values for the selected IMFs ranged between 0.6 and 0.96 for all seizures among all epilepsy patients. A threshold of the 75^th^ percentile of the *PLV* values in the *L* ***×*** *1* array was found to consistently result in pairs of channels with *PLV* greater than 0.6 for all seizures. It should be noted that pairs of channels with higher *PLV* values contribute more to the high phase-synchronization at the network level.
2. A list was formed with all pairs of channels with *PLV* values exceeding the threshold.
3. The number of times a channel appears in the list (as part of a pair) was defined as its degree of connection. The degree of connection of a channel is indicative of its effectiveness in controlling the network. That is, a node with a high degree of connection would entrain a larger portion of the network resulting in a significant increase in the network phase-synchrony [39]-[41].
4. Channels that appear at least twice (degree of connection greater than one) were considered candidate nodes for DBS. A channel with degree of connections equal to 0 is an isolated node and a channel with degree of connection equal to 1 is an end node. Both of these cases possess minimal potential for driving an epileptic network into a hyper-synchronization.
5. Channels or nodes that were considered as candidates and consistently appeared among all seizures of a patient were selected as final locations of stimulation.

### H. Frequency Selection for DBS

The phase of each IMF was measured using the Hilbert transform for each 1-s segment for the entire 10-s termination window. The frequency of each IMF was calculated by taking the time derivative of its instantaneous phases over each 1-s data segment. Finally, the average frequency over the 10-s termination window for each selected node, in the previous section, was measured as the stimulation frequency for that specific channel. The variation in frequency values across the 10-s termination window for any channel was less than 3%.

## III. Experimental Results

All seizures from the three epilepsy patients were analyzed using the proposed methodology. To deal with the edge effect problem associated with the data segmentation during the NA-MEMD process, 50 samples from neighboring segments were incorporated to both ends of each 1-s segment. However, after executing the NA-MEMD, only the IMFs corresponding to the original data segment were retained. Furthermore, although all IMFs were considered for the phase-synchrony analysis, only IMFs with frequency above 4 Hz were included in the subsequent analysis to determine the stimulation parameters. The IMFs with frequency below 4 Hz did not show any changes in their phase-connectivity with respect to seizures.

### A. Phase-Connectivity Analysis to Select DBS Parameters in Patient-1

Network phase-synchrony and phase-connectivity analysis were performed on seizures recorded in Patient-1 with TLE in order to determine stimulation frequency and locations. Fig. 4(a) illustrates the anatomical position of depth electrodes in Patient-1. Fig. 4(b) shows a 3-min iEEG signal, containing an epileptic seizure, used for this analysis. The green dashed lines indicate seizure onset and termination that were clinically determined by treating physicians. Temporal changes in the normalized *meanλ*_*1:60%*_ variable, which reflects the evolution of network phase-synchrony, is illustrated in Fig. 4(c). The normalized *meanλ*_*1:60%*_ value started to increase from about 15-s preceding seizure onset up to almost 30-s after it, which indicates a phase desynchronization among oscillators within this period. However, the synchrony level started to gradually increase toward seizure offset and reached its maximum level at seizure termination. This result suggests that the epileptic network is maximally synchronized at this point. Table II lists the *TM-PLV* values for all IMFs above the cut-off frequency of 4 Hz for all three patients. The IMF5 was selected as the bandwidth of interest, because it possessed the highest *TM-PLV* value. It should be noted that IMF5 consistently had the maximum *TM-PLV* value among all seizures in this patient.

**TABLE II.** *TM-PLV* VALUES AT TERMINATION WINDOW IN DIFFERENT IMFS AMONG ALL PATIENTS

| Subject | TM-PLV |  |  |  |  |  |
| --- | --- | --- | --- | --- | --- | --- |
|  | IMF 1 | IMF 2 | IMF 3 | IMF 4 | IMF 5 | IMF 6 |
| 1 | 0.31 | 0.28 | 0.26 | 0.38 | 0.75 | 0.35 |
| 2 | 0.48 | 0.69 | 0.41 | 0.43 | 0.48 | 0.45 |
| 3 | 0.73 | 0.93 | 0.80 | 0.62 | 0.34 | 0.53 |

**Fig. 4.**
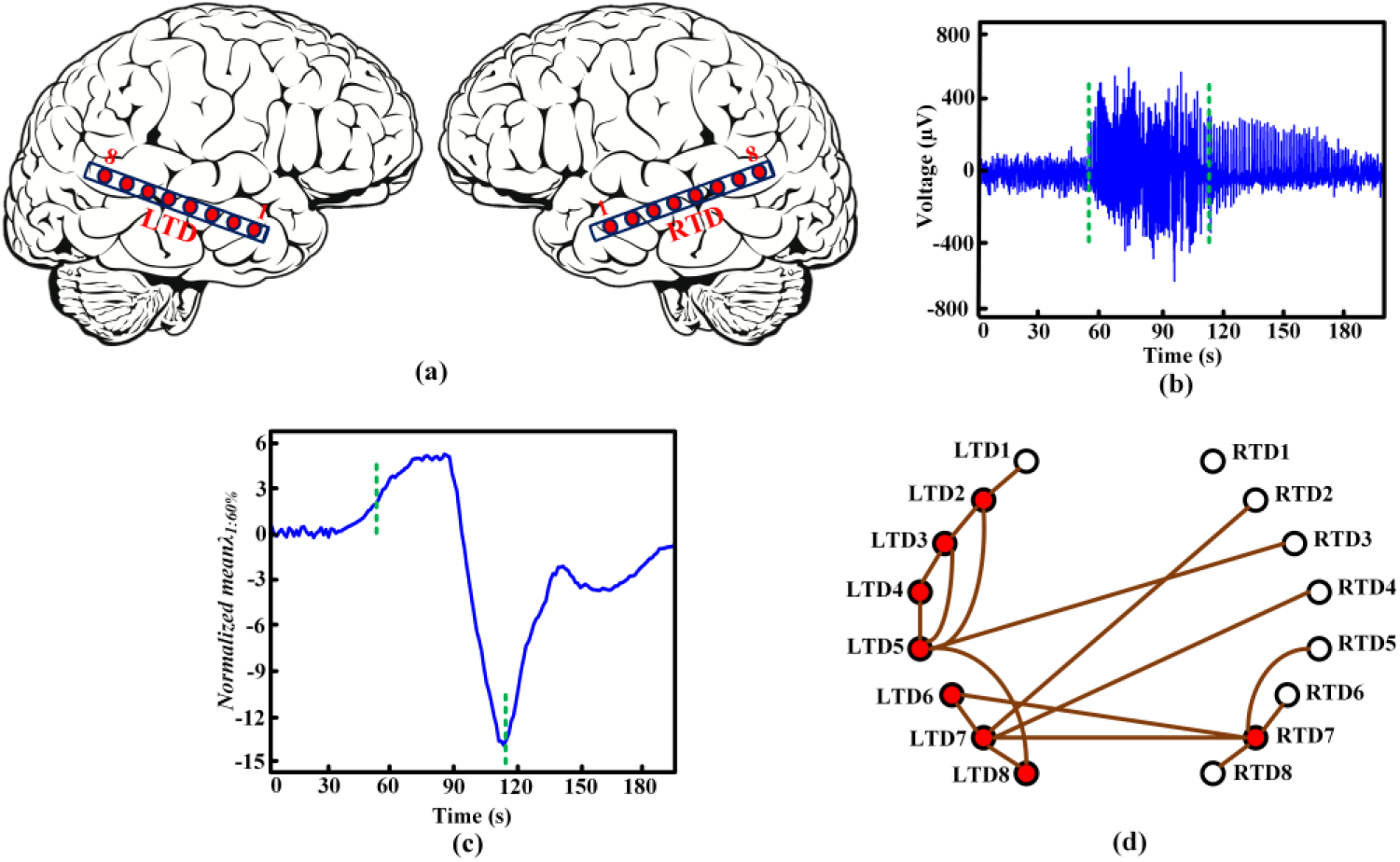
Phase-synchrony and phase-connectivity analysis for selecting DBS locations and frequency in Patient-1 with TLE. (a) Anatomical position of depth electrodes. (b) 3-min, iEEG recording containing a seizure. The green dashed lines denote seizure onset and offset. (c) Temporal changes of the normalized *meanλ*_*1:60%*_ value, after smoothing with a 5-s moving average filter, for IMFs extracted from all channels. An increase/decrease in the value of the normalized *meanλ*_*1:60%*_ corresponds to a decrease/increase in the phase-synchrony level, respectively. (d) Phase-connectivity analysis for the selected IMF, (IMF5). The brown lines drawn between pairs of channels indicate the ones with *PLV* value above the 75^th^ percentile threshold. The red-filled circles are the candidate nodes for DBS.

Fig. 4(d) displays the connections between unique pairs of channels for the selected IMF, (IMF5). The circles denote the recording sites on the depth electrodes implanted in Patient-1. The brown lines drawn between pairs of channels indicate the ones with *PLV* value above the 75^th^ percentile threshold. The red-filled circles are the candidate nodes for DBS. This analysis was performed on all recorded seizures for Patient-1. The final nodes, candidate nodes that appeared in all seizures as described in methods section part G, for this patient were LTD5, LTD7 and RTD7. It should be noted that the final nodes for DBS correlated well with the channels possessing the highest degree of connections. These channels appeared to be hubs and thereby are well suited for synchronizing the epileptic network. All of these channels were located in the hippocampus and the measured frequency of them within the termination window was approximately 15 Hz. The measured frequency was the same among all seizures for this patient and was chosen as the proposed stimulation frequency.

### B. Phase-Connectivity Analysis to Select DBS Parameters in Patient-2

The same analysis described in the previous section was carried out for Patient-2. Fig. 5 displays the phase-synchrony and phase-connectivity dynamics for an iEEG data containing an epileptic seizure in Patient-2 with ETE. Fig. 5(a) shows the anatomical position of the depth electrodes in the patient. A 4-min iEEG signal utilized for the phase-synchrony and phase-connectivity analysis, is shown in Fig. 5(b). Green dashed lines indicate seizure onset and offset. Fig. 5(c) exhibits the normalized *meanλ*_*1:60%*_ obtained from the phase-synchrony analysis. The phase-synchrony pattern was similar to the one observed in Patient-1. The network achieved the maximum phase-synchrony level at seizure termination. The bandwidth of interest was measured to be the second IMF (IMF2) as it had the highest *TM-PLV* value, which is reported in Table II. The selected IMF was the same among all seizures analyzed in this patient. Fig. 5(d) displays the candidate nodes for DBS for a recorded seizure in Patient-2 using the same procedure applied to Patient-1. The final nodes for DBS were identified as LAD2 and LAD3. The final nodes for DBS overlapped well with the obtained candidate nodes with the highest degree of connections in the network. These sites appeared to be hubs in the network and further supported selecting these channels as simulation sites for DBS. Both of these channels were located in the amygdala and the measured frequency of them within the termination window was about 90 Hz. The calculated frequency was the same among all seizures for this patient and was selected as the proposed stimulation frequency.

**Fig. 5.**
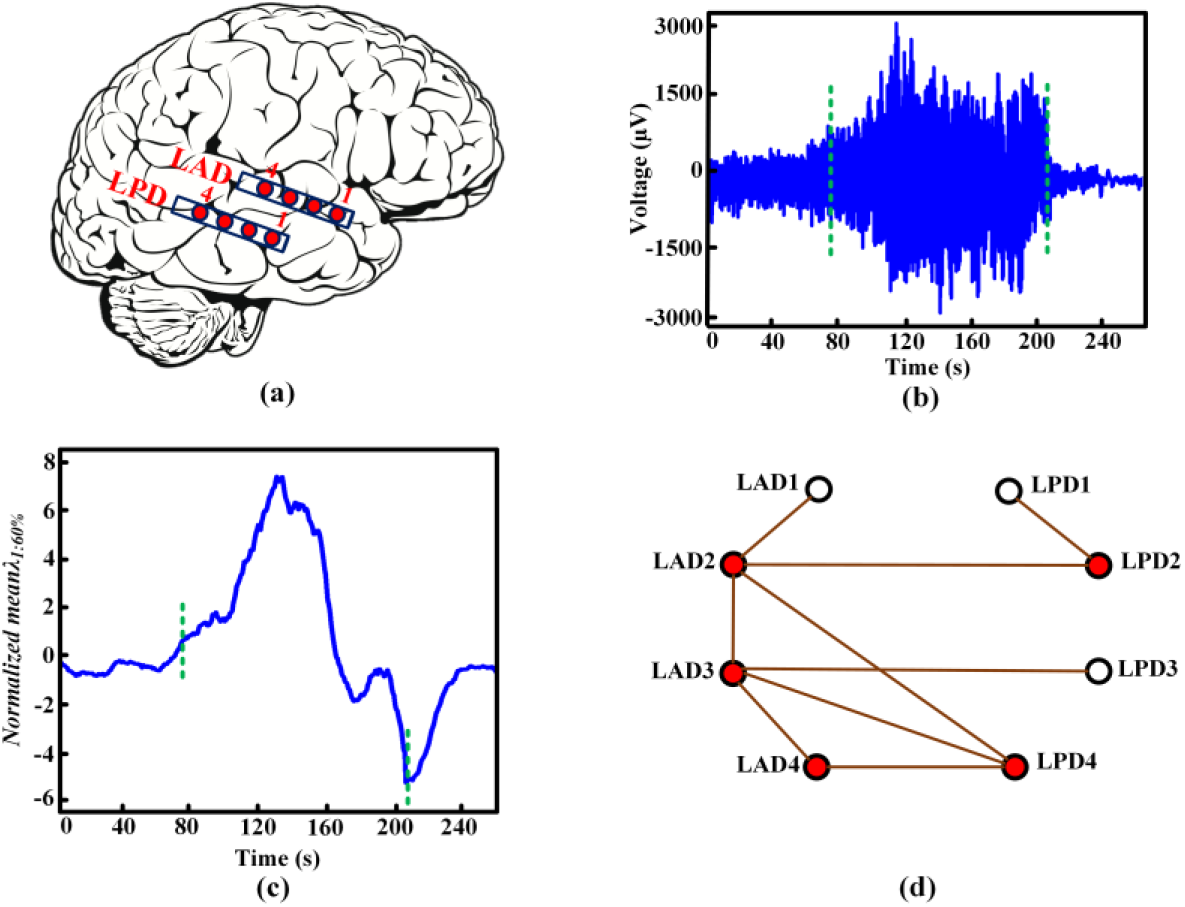
Phase-synchrony and phase-connectivity analysis for selecting DBS locations and frequency in Patient-2 with ETE. (a) Anatomical position of depth electrodes. (b) A 4-min iEEG recording containing a seizure. The green dashed lines denote seizure onset and offset. (c) Temporal changes of the normalized *meanλ*_*1:60%*_ value, after smoothing with a 5-s moving average filter, for IMFs extracted from all channels. An increase/decrease in the value of the normalized *meanλ*_*1:60%*_ corresponds to a decrease/increase in the phase-synchrony level, respectively. (d) Phase-connectivity analysis for the selected IMF, (IMF2). The brown lines drawn between pairs of channels indicate the ones with *PLV* value above the 75^th^ percentile threshold. The red-filled circles are the candidate nodes for DBS.

### C. Phase-Connectivity Analysis for the Selection of DBS Parameters in Patient-3

Fig. 6 exhibits the phase-synchrony and phase-connectivity dynamics for an iEEG signal containing a seizure in Patient-3 with ETE. Fig. 6(a) illustrates the anatomical position of the depth electrodes in this patient. Fig. 6(b) shows a 4-min iEEG signal utilized for this analysis. Green dashed lines denote seizure onset and offset. The normalized *meanλ*_*1:60%*_ variable obtained from the phase-synchrony analysis is shown in Fig. 6(c). A phase desynchronization was observed from ictal onset to mid-ictal. However, the synchrony level started to gradually increase from mid-ictal towards ictal offset and achieved its maximum level at seizure termination. As shown in Table II, the second IMF (IMF2) was obtained as the bandwidth of interest as it had the highest *TM-PLV* value among the other extracted IMFs. Moreover, IMF2 was consistently obtained as the IMF with the maximum *TM-PLV* value among all seizures analyzed in this patient. Fig. 6(d) displays the candidate nodes for DBS for one of the recorded seizures in Patient-3 using the analysis reported in the method section. For Patient-3, the final nodes for stimulation were identified as LAD3 and LPD2. The final nodes for DBS overlapped well with the candidate nodes with high degree of connections. Besides, these channels appeared to be hubs in the network with respect to their degree of connections. One of the channels was located in the amygdala and the other was located in the hippocampus. The frequency of these nodes within the termination window was obtained to be approximately 90 Hz. This frequency was the same among all seizures recorded in this patient, and thereby was selected as the proposed stimulation frequency.

**Fig. 6.**
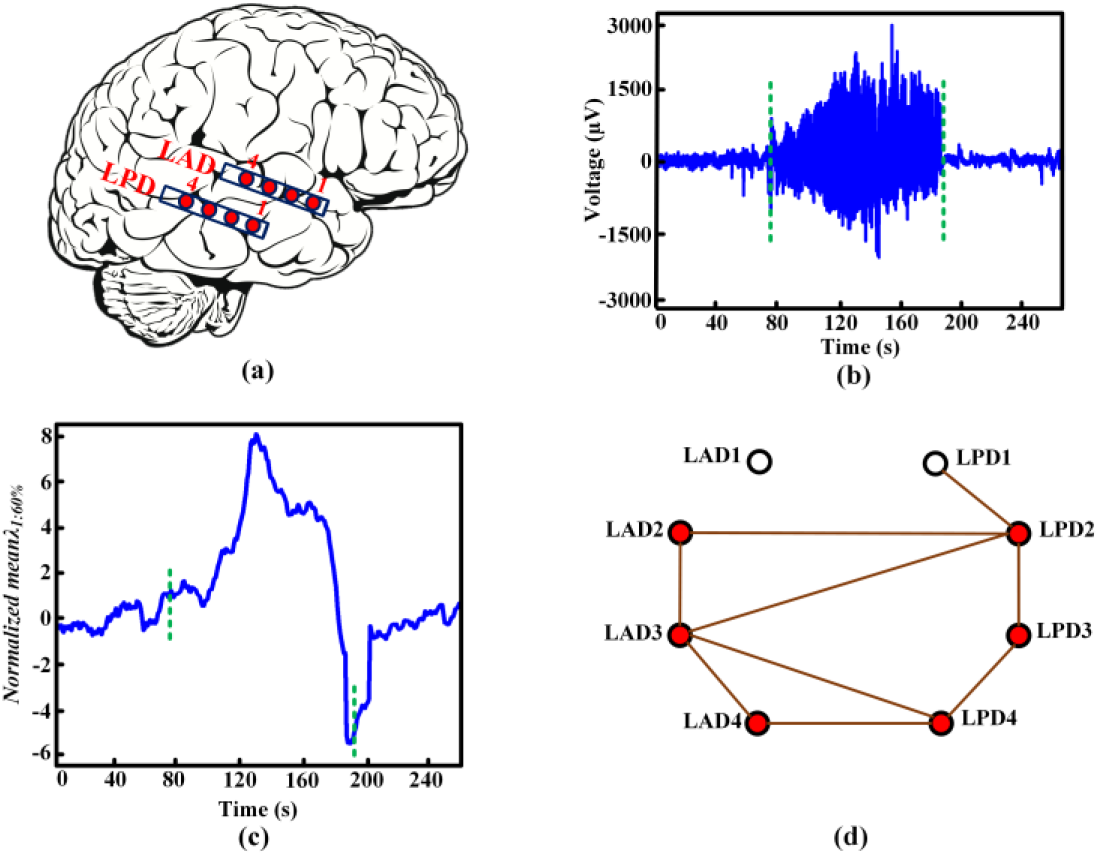
Phase-synchrony and phase-connectivity analysis for selecting DBS locations and frequency in Patient-3 with ETE. (a) Anatomical position of depth electrodes. (b) A 4-min iEEG recording containing a seizure. The green dashed lines denote seizure onset and offset. (c) Temporal changes of the normalized *meanλ*_*1:60%*_ value, after smoothing with a 5-s moving average filter, for IMFs extracted from all channels. An increase/decrease in the value of the normalized *meanλ*_*1:60%*_ corresponds to a decrease/increase in the phase-synchrony level, respectively. (d) Phase-connectivity analysis for the selected IMF, (IMF2). The brown lines drawn between pairs of channels indicate the ones with *PLV* value above the 75^th^ percentile threshold. The red-filled circles are the candidate nodes for DBS.

## IV. Discussion and Conclusion

In this study, an adaptive non-linear analytical methodology was proposed that merges both phase-synchrony and phase-connectivity analysis in order to extract stimulation frequency and locations that match the endogenous brain dynamics of patients as seizures naturally terminate. This method provides an algorithm for tuning DBS parameters in order to develop a DBS protocol that can be delivered upon detection of subsequent seizures in an effort to terminate them immediately. Applying DBS using the proposed closed-loop approach could potentially improve the clinical efficacy of the therapy as shown in our previous study performed in a freely moving chronic rat model of limbic epilepsy [22]. The analysis proposed in this study are divided into two major components; namely, (1) Phase-synchrony analysis in order to determine how network synchrony changes as seizures evolve, and (2) Phase-connectivity analysis in order to select IMF (bandwidth of interest), channels (locations of stimulation), and finally frequency of electrical stimulation. These parameters were extracted to match the maximum level of phase-synchrony observed at seizure termination in order to reproduce hyper-synchrony.

The first step in the phase-synchrony analysis was the decomposition of wideband electrophysiological signals into a set of finite, narrow-band neuronal oscillators using NA-MEMD. The instantaneous phases of the oscillators or IMFs were then calculated using the Hilbert transform. Next, the eigenvalue decomposition of the square, bivariate mean-phase coherence matrix, *R*_*N×N*_, was performed to access the phase-synchrony dynamics between IMFs. The normalized mean value of the first 60% lower-index eigenvalues, *meanλ*_*1:60%*_, was measured and reported for each second to determine changes in the network’s synchrony level as seizures evolve. In all patients, a decrease in phase-synchrony was observed from seizure onset to mid-ictal. This finding is consistent with what has been reported by other researchers [42], [43]. The phase-desynchronization period was followed by a gradual increase in phase-synchronization level as seizure progressed and reached its maximum level at seizure termination. The hyper-synchrony at seizure termination provided the basis for the phase-connectivity analysis, and is consistent with recent studies using human and rat epileptic seizures [42], [44], [45]. The phase-connectivity analysis used bivariate phase-locking value, *PLV*, to locate the neuronal oscillators that contribute the most to the hyper-synchronization observed at seizure offset. This was achieved by finding the neuronal oscillators that were highly synchronized in the 10-s window leading up to seizure termination.

Both the obtained frequency and locations of stimulation representing the hyper-synchronization at termination window varied among the patients analyzed in this study. It should be noted that the extracted stimulation frequency and locations were stable among all seizures in all patients. Patient-1 with TLE had identified locations in the hippocampus while the other two patients with ETE had at least one of their site outside of the hippocampus. The frequency of stimulation obtained in Patient-1 was lower than the other two patients. Patient-1 had bilateral hippocampal sclerosis while the other two patients possessed no lesions. These results suggest that the underlying pathology may contribute to the variance in frequency and locations of stimulation obtained in epilepsy patients. Most DBS therapy using available FDA-approved devices utilize pre-determined stimulation parameters such as stimulation frequency and locations for a cohort of patients. In clinical studies, patients showing more than 50% reduction in seizure frequency in comparison to baseline are labelled as responders and those that do not, are categorized as non-responders. The therapeutic DBS protocol described in this study could potentially increase the responder’s rate using current DBS devices by accounting for patient’s specific brain dynamics when determining stimulation parameters. A recent study that investigated hippocampal stimulation in patients with drug-resistant mesial temporal lobe epilepsy (MTLE) found individual variation in the optimal DBS frequency [46]. Low frequency hippocampal stimulation was effective (more than 50% reduction in seizure frequency) in MTLE patients with hippocampal sclerosis, while it was ineffective (∼20% reduction in seizure frequency) in patients with no lesions in the hippocampus. Switching stimulation frequency in a patient with no lesion to a high frequency resulted in greater reduction (up to 55%) in seizure frequency relative to the baseline level. Furthermore, several studies have reported the efficacy of high frequency DBS in reducing seizure frequency for epilepsy patients [47]-[49]. On the other hand, many other studies have suggested that low frequency stimulation is more efficacious in reducing seizure frequency [50]-[52]. Computational results of our analysis also supported the difference in stimulation frequency and locations that matched the hyper-synchrony at seizures termination among epilepsy patients.

In addition to providing stimulation parameters based on patient’s brain dynamics, another important feature of closed-loop stimulation is the ability to deliver electrical stimulation in response to seizure detection. In all seizures recorded from patients, there is a decrease in the network’s phase-synchrony level a few seconds before seizure onset. This decrease in phase-synchrony may be utilized as a potential trigger in implantable neuromodulation devices for applying electrical stimulation in order to terminate epileptic seizures.

The main aim of this study was to extract DBS stimulation frequency and locations that drive the epileptic network into hyper-synchrony at natural seizure termination. This DBS protocol can be delivered upon seizure detection to artificially revert the phase desynchronization dynamics observed around seizure onset in order to terminate it immediately.

This analysis is adaptive and takes into consideration the endogenous brain dynamics of patients that could potentially leads to personalizing and optimizing DBS therapy for them. Future studies will focus on applying the proposed method to a higher number of epilepsy patients and exploring its clinical efficacy through DBS using the patient-specific stimulation parameters.

## Data Availability

The data is available and public in IEEG portal created by Mayo Clinic.
Reference of the portal and data is cited in the paper. We only used clinical data and we never performed any clinical study.
Clinical data was adopted from IEEG portal:
https://www.ieeg.org

